# Automated Detection of Motor Speech Disorders and Subtype Classification

**DOI:** 10.64898/2026.07.16.26358268

**Authors:** Fenqi Wang, Rene L. Utianski, Leland R. Barnard, John L. Stricker, Heather M. Clark, Gabriela F. Meade, David T. Jones, Jennifer L. Whitwell, Keith A. Josephs, Joseph R. Duffy, Hugo Botha

## Abstract

Motor speech disorders (MSDs) are early markers of neurological disease, but expert perceptual analysis is rarely available outside specialized centers. Automated speech analysis offers a scalable alternative, yet prior studies have not systematically compared modeling approaches or assessed clinically relevant metrics in independent datasets. This study compared static acoustic features, articulatory informed Phonet features, and self-supervised pretrained models for binary and multi label MSD classification.

We trained and evaluated models on 583 speech samples using speaker level splits. Baseline models included logistic regression and Gated Recurrent Units (GRUs) trained on eGeMAPS and MFCCs. We extracted three types of Phonet derived features and evaluated pretrained HuBERT and SSAST models in frozen, partially fine-tuned, and fully fine-tuned configurations. Binary classification distinguished MSDs from controls, while multi label classification identified six MSD subtypes. Models were assessed using validation AUC, and cut points were tested on two independent datasets.

Pretrained and Phonet based models substantially outperformed static acoustic features. In binary classification, HuBERT achieved the highest AUC (0.95), while compact Phonet derived GRUs achieved comparable performance (up to 0.94). These models generalized well to independent datasets, maintaining high sensitivity (0.94) and specificity (0.97). In multi label classification, Phonet models achieved the highest macro average AUC (0.86), but threshold-based subtype performance declined on unseen data.

Automated MSD detection is feasible and clinically promising. Binary classification generalized well, whereas multi label classification showed limited threshold stability across datasets.

## Introduction

Neurological diseases are among the most common and debilitating conditions worldwide, often leading to chronic disability and a diminished quality of life (Feigin et al., 2019). Early and accurate diagnosis is essential for optimizing treatment and improving long-term outcomes (Espay et al., 2017; Postuma et al., 2015). Yet many neurological disorders are either misdiagnosed or diagnosed too late, in part due to the subtlety and overlap of early clinical signs (Abrahams et al., 2014; Dang et al., 2021; Litvan et al., 2003).

Speech changes are an early and distinctive manifestation of several neurological diseases (Duffy, 2008, 2020). Motor speech disorders (MSDs), including the dysarthrias and apraxia of speech, reflect damage to different neural circuits involved in speech planning, programming, and execution (Shipley & McAfee, 1992). Because the type and severity of speech abnormality often correspond to specific neurological loci, perceptual analysis of speech can offer valuable diagnostic information (Duffy et al., 2014; Melle & Gallego, 2012). However, expert perceptual characterization of speech is rarely available outside specialized centers (Duffy, 2020; Kent, 2004), as it requires extensive training, subjective judgment, and time (Bunton et al., 2007; Patel et al., 2013). As a result, many patients go without a thorough speech-based evaluation, leading to diagnostic delays and missed opportunities for early intervention (Dang et al., 2021; Liss et al., 2009).

To address the limitations of expert-dependent evaluations, research has increasingly turned to automated acoustic analyses that provide objective, scalable, and reproducible measures. Most studies have relied on static features, such as Mel-frequency cepstral coefficients (MFCCs) or extended Geneva Minimalistic Acoustic Parameter Set (eGeMAPS) combined with machine learning classifiers. More recently, deep learning approaches have surpassed traditional machine learning for speech processing, but small clinical datasets preclude their widespread use. Self-supervised pretrained models such as HuBERT and SSAST offer a solution by learning general-purpose speech representations that can be applied to small datasets and adapted for specific tasks. Early applications suggest these models can capture neuropathological changes in neurological speech (Cadet et al., 2024; Grant & West, 2024), though they are computationally intensive and not optimized for the articulatory deficits that pervade MSDs.

An alternative is Phonet, a model trained with an explicit articulatory objective. Phonet extracts posterior probabilities (Vásquez-Correa et al., 2019), encoding articulatory properties such as manner and place of articulation, making its features clinically interpretable. Prior work shows these features correlate strongly with perceptual ratings and detect pathological patterns across conditions, such as Parkinson’s disease and apraxia of speech (Cernak et al., 2017), positioning Phonet as a promising alternative to large-scale pretrained systems.

Despite these advances, several important gaps remain. No study has systematically compared these approaches within a unified framework (Eyben et al., 2016; Gong et al., 2022; Hsu et al., 2021; Javanmardi et al., 2024; Little et al., 2009; Rowe & Green, 2019), assessed clinically relevant metrics requiring cut-points and categorical decisions as to the presence or absence of disease, or evaluated generalization to independent datasets. Most prior studies focus on the detection of simple MSD presence, one or two MSDs, whereas clinicians face a broader differential diagnosis in clinical practice.

This study addresses these gaps by comparing static acoustic features, articulatory- informed Phonet features, and large pretrained models for both binary classification (presence vs. absence of MSD) and multi-label classification (six MSD subtypes). We define cut points in a validation set and test generalization in independent datasets. We hypothesize that binary classification models will generalize well, while multilabel models will be less robust. Our goal is to evaluate the potential of these tools to aid clinicians in detecting MSDs.

## Methods

### Datasets

Speech samples were obtained from patients repeating a single instance of the sentence ‘*My physician wrote out a prescription*’ following a spoken model as part of a recording session that included a standard reading passage, picture description, word and sentence repetition, sustained vowel, speech alternating motion rate (rapid repetition of “puh”), and speech sequential motion rate (rapid repetition of “puh tuh kuh”). Some participants completed additional recording sessions over time at ∼4 monthly or yearly intervals. For each recording session, a participant could be labeled with one or more of the following six MSDs: flaccid dysarthria, spastic dysarthria, ataxic dysarthria, hypokinetic dysarthria, hyperkinetic dysarthria, and apraxia of speech.

These MSD diagnoses were assigned by a board-certified speech-language pathologists based on review of all the tasks in each recording session, but not across sessions, meaning a participant may have received different MSD diagnoses across recording sessions. All adult English-speaking patients seen in the department neurology were invited to participate in a study where they complete a remote speech-language exam, with no additional inclusion or exclusion criteria.

The overall dataset consisted of 583 recording sessions from 456 speakers, including 259 recordings from 234 individuals not labelled with any MSD and 324 recordings from 229 individuals with at least one subtype of MSD. The majority of speakers were White (93.4%) and non-Hispanic or Latino (94.5%).

The dataset was stratified into training and validation sets using speaker-level splitting to ensure no speaker appeared in both sets. The same data splits were maintained across all experiments to ensure fair model comparisons. The training set included 262 recordings with at least one MSD subtype across 183 speakers and 209 recordings from 187 speakers without an MSD. The validation set included 62 recordings with at least one MSD subtype across 45 speakers and 50 recordings from 47 speakers without an MSD.

To test the best model(s) on a true hold out set, we also identified recordings of the same sentence from two independent datasets at Mayo Clinic, MN with no overlapping speakers with the main dataset (see details in the Supplement). As such, these test sets are not only completely independent from our discovery dataset, but they were also recorded in different settings without the standardized prompt and example used in our remote capture battery, and the labels are from full clinical or full research evaluations rather than being based on the brief recordings themselves. This dataset represents a more realistic, albeit far more difficult, test of model generalization.

The distribution of MSD subtypes across datasets is presented in Table 1, whereas more detailed breakdowns of the co-occurrence of MSDs and distribution of age and sex across datasets are included in the Supplemental Data (see Supplemental Figures 1-3).

**Table 1.** Distribution of MSD Subtypes in the Training, Validation, and Test Datasets.

|  | None | Flaccid | Spastic | Ataxic | Hypokinetic | Hyperkinetic | Apraxia |
| --- | --- | --- | --- | --- | --- | --- | --- |
| **Training** |  |  |  |  |  |  |  |
| Participants* | 187 | 40 | 50 | 37 | 33 | 48 | 18 |
| Recordings | 209 | 53 | 66 | 54 | 39 | 58 | 44 |
| **Validation** |  |  |  |  |  |  |  |
| Participants* | 47 | 11 | 12 | 11 | 7 | 10 | 5 |
| Recordings | 50 | 14 | 16 | 15 | 8 | 10 | 11 |
| **Test (NRG)** |  |  |  |  |  |  |  |
| Participants* | 49 | 0 | 4 | 5 | 2 | 0 | 23 |
| **Test (JRD)** |  |  |  |  |  |  |  |
| Participants* | 11 | 7 | 15 | 22 | 17 | 12 | 9 |

*Note: \*The sum of number of participants in the row exceeds the number in the dataset because each participant can have more than one MSD.*

### Feature Representations and Model Families

We evaluated four main families of models, each reflecting a different way of representing and analyzing speech. These ranged from simple, handcrafted acoustic features to large pretrained neural networks that learn their own representations.

The first model family used logistic regression (LG) with static acoustic features, which summarize entire recordings into fixed-length vectors. Two feature sets were tested. The first, eGeMAPS (model 1a),(Eyben et al., 2016) was extracted with the openSMILE toolkit and consists of widely used summary statistics of basic voice characteristics such as pitch, spectral shape, and voice quality. The second, 13 MFCCs (model 1b) were extracted per frame of each recording and then summarized by their mean and standard deviation, producing a 26-dimensional vector. In both cases, recordings were standardized by resampling to 16 kHz, trimming to a max of 10 seconds, and extracting one feature vector per participant.

The second model family extended this approach by adding temporal modeling with gated recurrent units (GRUs), a type of recurrent neural network ideal for capturing sequential dynamics in speech while keeping the parameter count manageable enough to apply to small datasets. Instead of using only summary features, these models operated directly on frame-level representations. Model 2a used low-level descriptors from the eGeMAPS set (e.g., pitch and spectral measures calculated every 10 ms), while model 2b used framewise MFCCs. In both cases, the GRUs learned how patterns evolved over time, providing richer representations than static features alone.

The third model family used articulatory-informed features from Phonet with GRUs. Phonet (Vásquez-Correa et al., 2019), a hierarchical recurrent neural network, is designed to capture the articulatory structure of speech. Input speech was segmented into short frames (25 ms with 10 ms overlap), converted into mel filterbank representations, and passed through recurrent layers that produced posterior probabilities for phonemes and phonological features. To adapt the model for English, we retrained Phonet on the LibriSpeech corpus (360 hours of English speech recordings). Using Phonet, we extracted three types of features: (1) posteriorgrams, representing frame-level probabilities of phonemes (model 3a); (2) phonological class embeddings, taken either at the frame level (model 3b_1) or mean-pooled across time (model 3b_2); and (3) phoneme embeddings, extracted either at the frame level (model 3c_1) or mean-pooled across time (model 3c_2). These features were then used as input to GRU classifiers, while the Phonet model itself remained fixed.

The fourth model family consisted of large pretrained self-supervised transformer models, HuBERT (Hsu et al., 2021) and SSAST (Gong et al., 2022), which learn speech representations directly from raw audio and spectrograms, respectively. Each was tested in three configurations: using the pretrained model HuBERT or SSAST as a frozen feature extractor with only a classifier trained on top (models 4a and 4d, respectively), partially fine-tuning the top layers (models 4b and 4e, respectively), or fully fine-tuning all layers (models 4c and 4f, respectively) (Wiepert et al., 2024).

### Classification Tasks and Training Procedures

We evaluated two types of classification tasks. The first was a binary task distinguishing recordings with any MSD from controls. The second was a multi-label task, in which each of the six MSD subtypes was treated as a separate binary label.

Training procedures were tailored to each model family while maintaining identical data splits to ensure comparability. Logistic regression models (models 1a and 1b) were trained with L2 regularization, with model complexity tuned on the validation set. GRU-based classifiers (models 2a, 2b, and 3a–3c) were used for models incorporating temporal dynamics, and pretrained self-supervised models HuBERT (models 4a–4c) and SSAST (models 4d–4f) were evaluated in frozen, partially fine-tuned, and fully fine-tuned configurations. All models were optimized using standard procedures appropriate to their architecture. To ensure robustness, models were trained under multiple random initializations and learning rates, with early stopping based on validation performance. Detailed architecture specifications, optimization parameters, and hyperparameter search procedures are provided in the Supplement.

### Evaluation Metrics and Model Selection

Model performance was assessed using the area under the receiver-operating characteristic curve (AUC). For binary classification, we computed overall validation AUC for each model, while for multi-label classification we computed the macro-average AUC across all six MSD subtypes as well as subtype-level AUCs. Detailed model selection procedures and subtype-specific comparisons are provided in the Supplement.

After the most promising binary model was identified, we assessed validation performance in the following validation data subsets to evaluate bias: stratified by gender, stratified by age split at the median, stratified by the intersection of gender and age tertiles. The small numbers of each label in the validation set made slices of the data unfeasible for the multilabel models.

To translate the model’s continuous output predictions into actionable classifications, we explored two approaches for setting decision thresholds. The first was based on the receiver-operating characteristic (ROC) curve, but with the penalty for false negatives set at 2 x that of false positives, in keeping with the intended use of the model as a screening tool. Given the limited sample size of positive cases in the multilabel scenario it is likely that ROC based cut points will be brittle. As such, for the second method we avoided empirical ROC optimization for threshold selection. Instead, we established reference intervals based on the healthy validation cohort, defining the cut point as the 95th percentile (*Z*=1.645) of the healthy distribution, similar to what might be done for neuropsychological tests. For each label both cut-points were assessed in the validation cohort and the best approach was selected based on permutation testing of sensitivity, specificity, and balanced accuracy, as well as visual inspection of the cut-points relative to the predicted probability distributions for each class. These thresholds were then used to report performance in the validation set and subsequently applied to the test set for final evaluation.

### Standard Protocol Approvals and Patient Consents

This study was approved by the Institutional Review Board (IRB) of Mayo Clinic (IRB No. 22-002430). Written informed consent was obtained from all participants prior to enrollment, including consent for the use of their audio recordings for research purposes. The study was conducted in accordance with the ethical principles of the Declaration of Helsinki. All audio recordings were anonymized prior to analysis, and all study procedures complied with applicable data protection and privacy regulations.

### Data Availability

Anonymized feature data and scripts used for model training and evaluation will be shared with qualified researchers upon request and under a data use agreement.

## Results

### Binary Classification

In the binary classification task, where the goal was to distinguish between individuals with and without an MSD, performance varied substantially across model families (see Figure 1). Baseline logistic regression models using static acoustic features were the weakest overall. The eGeMAPS-based model (model 1a) achieved a validation AUC of 0.857, while the MFCC-based model (model 1b) reached only 0.669. Incorporating temporal modeling with GRUs slightly improved results for eGeMAPS (model 2a: 0.833) but not for MFCCs (model 2b: 0.738).

**Figure 1.**
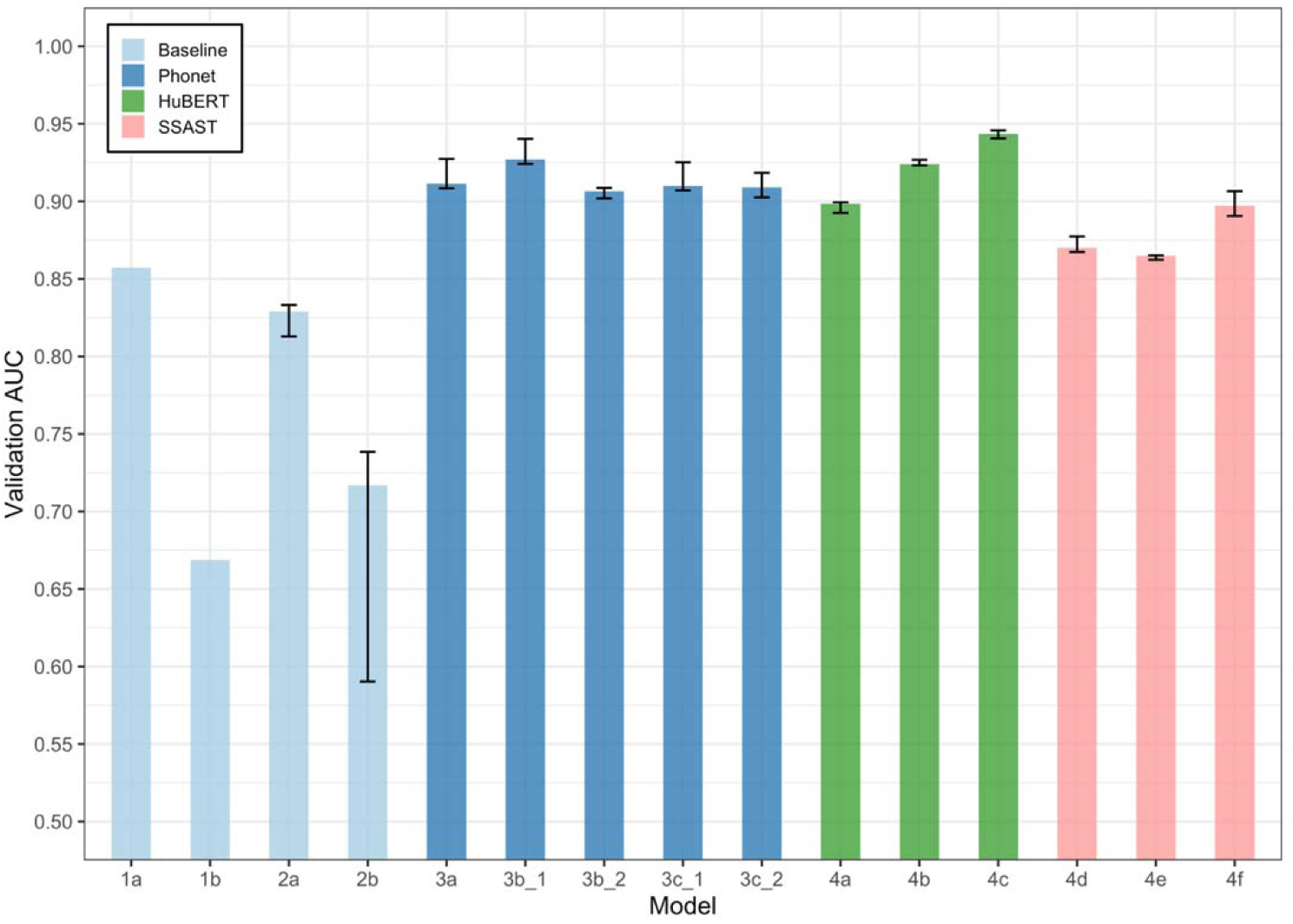
Binary classification performance across models. Bars show median validation AUCs, and error bars indicate the range (minimum to maximum) across seeds.

Phonet-derived models consistently outperformed the static acoustic baselines. Posteriorgrams (model 3a) reached an AUC of 0.927, while phonological class embeddings achieved 0.940 (model 3b_1) and 0.909 (model 3b_2). Phoneme embeddings also performed strongly, with model 3c_1 reaching 0.925 and model 3c_2 0.918. These compact models approached the performance of much larger pretrained architectures such as HuBERT and SSAST.

Among the pretrained self-supervised models, HuBERT achieved the highest overall AUC, with full fine-tuning (model 4c) yielding 0.946. Partial fine-tuning (model 4b: 0.927) and frozen embeddings (model 4a: 0.899) also performed competitively. SSAST models were slightly weaker overall, with the best performance under full fine-tuning (model 4f: 0.907), followed by frozen (model 4d: 0.877) and partial fine-tuning (model 4e: 0.865).

### Multi-label Classification

In the multi-label classification task, where each of the six MSD subtypes was treated as an independent binary label, performance was lower overall compared to the binary task, reflecting the added difficulty of handling overlapping and co-occurring diagnoses (see Figure 2).

**Figure 2.**
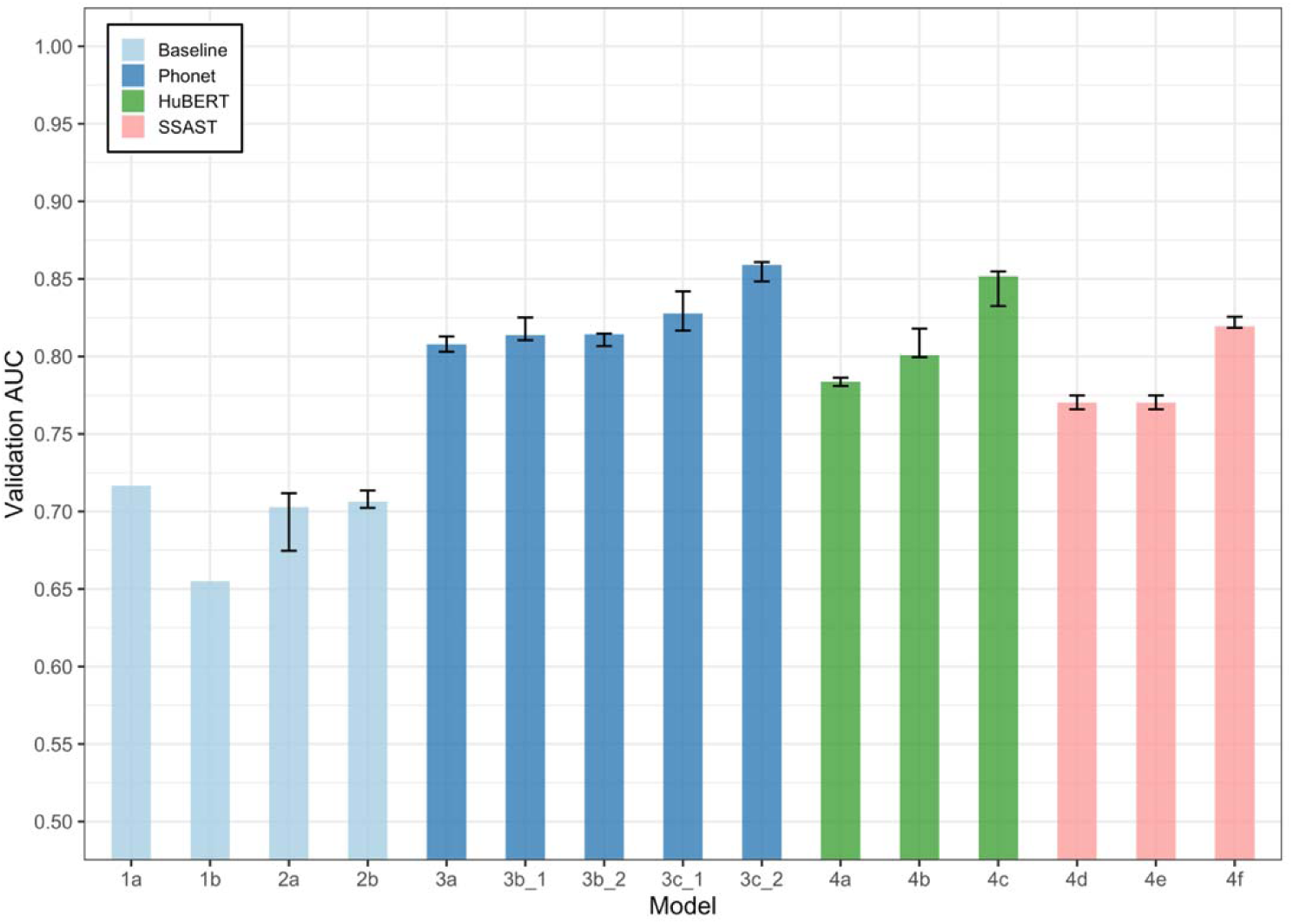
Multi-label classification performance across models. Bars show median validation AUCs, and error bars indicate the range (minimum to maximum) across seeds.

Baseline models performed the worst, with logistic regression on eGeMAPS (model 1a) achieving an AUC of 0.717 and MFCCs (model 1b) falling to 0.655. Adding temporal modeling via GRUs did not yield major improvements, with eGeMAPS-based GRUs (model 2a: 0.712) and MFCC-based GRUs (model 2b: 0.714) performing similarly.

Phonet-derived models demonstrated the strongest performance. Phoneme embeddings with mean pooling (model 3c_2) achieved the highest macro-average AUC overall at 0.861, followed by frame-level phoneme embeddings (model 3c_1: 0.842), phonological class embeddings (model 3b_1: 0.825; model 3b_2: 0.815), and posteriorgrams (model 3a: 0.813).

Among the pretrained self-supervised models, HuBERT again performed strongly, with full fine-tuning (model 4c) yielding 0.855. Its partially fine-tuned (model 4b: 0.818) and frozen (model 4a: 0.786) variants were less effective. SSAST models performed slightly lower, with the fully fine-tuned version (model 4f: 0.826) outperforming frozen (model 4d: 0.775) and partial fine-tuning (model 4e: 0.775).

### Subtype-wise Performance

We also examined subtype-specific AUCs using the best-performing configuration from each model family in the multi-label classification task (see Figure 3).

**Figure 3.**
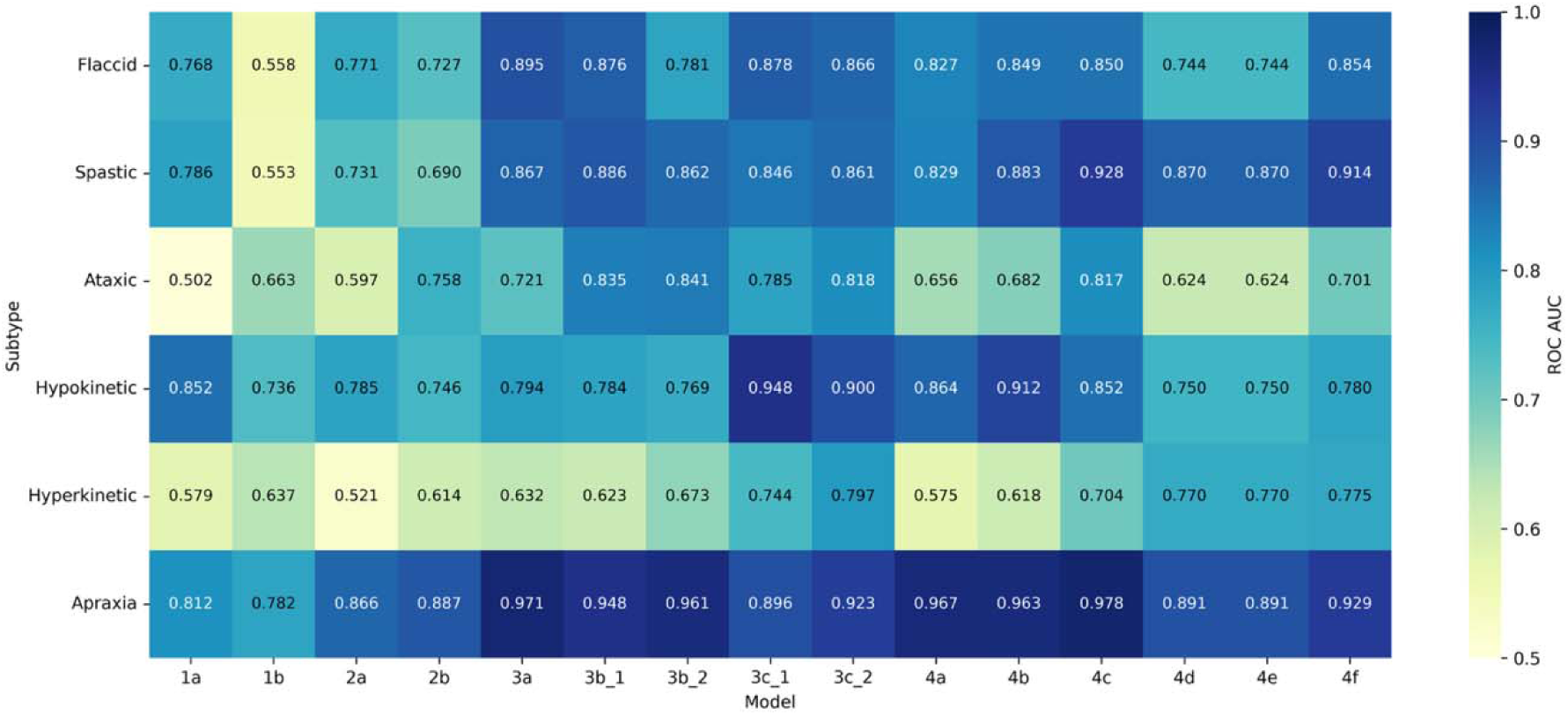
Subtype-specific validation AUCs for the best-performing configuration of each model family in the multi-label classification task.

The baseline acoustic models performed weakest overall. Model 1a reached 0.768 for flaccid and 0.812 for apraxia of speech but fell to 0.502 for ataxic and 0.579 for hyperkinetic dysarthria. Model 1b improved ataxic (0.663) and apraxia (0.782) but remained low for flaccid (0.558), spastic (0.553), and hyperkinetic (0.637). GRU-based baselines performed better, with model 2a scoring 0.866 for apraxia and 0.771 for flaccid but still only 0.597 for ataxic and 0.521 for hyperkinetic, while model 2b reached 0.887 for apraxia and 0.758 for ataxic but lagged on spastic (0.690) and hyperkinetic (0.614).

Phonet-derived models were consistently stronger. The best configuration, model 3c_2, achieved 0.923 for apraxia, 0.900 for hypokinetic, and 0.861 for spastic, with flaccid (0.866) and ataxic (0.818) also well captured. Hyperkinetic dysarthria remained the most difficult (0.797), though still higher than any baseline model. Pretrained models performed best in absolute terms but varied across subtypes. Model 4c achieved near-ceiling results for apraxia (0.978) and spastic (0.928), competitive scores for flaccid (0.850) and ataxic (0.817), but weaker for hypokinetic (0.852) and hyperkinetic (0.704). SSAST followed a similar pattern, with model 4f performing well for apraxia (0.929) and spastic (0.914) but showing lower values for hypokinetic (0.780) and hyperkinetic (0.776).

Across models, apraxia was the most reliably detected, with AUCs exceeding 0.78 in all families. Flaccid and spastic were also well identified, especially by Phonet (model 3c_2: 0.866 and 0.861) and HuBERT (model 4c: 0.850 and 0.928). Hypokinetic was well captured by Phonet (model 3c_1: 0.948) but less so by SSAST (model 4f: 0.780). Ataxic showed wide variability, from 0.502 in model 1a to 0.841 in model 3b_2. Hyperkinetic remained the most challenging subtype overall, with all models between 0.521 and 0.797.

Because hyperkinetic was consistently the hardest subtype to identify, we retrained models 3c_2, 4c, and 4f as binary classifiers (hyperkinetic vs. non-hyperkinetic) and compared them with their multi-label counterparts across seeds and learning rates (see Supplemental Figure 4). For Phonet (model 3c_2), binary performance fluctuated widely (0.314–0.800), while the multi-label version was consistently higher and more stable (0.713–0.808), suggesting benefits from the co-occurrence of subtypes. For HuBERT (model 4c), binary training was superior, producing AUCs between 0.638 and 0.749 compared to 0.503–0.704 for multi-label, suggesting reduced interference when isolating hyperkinetic dysarthria. SSAST (model 4f) showed a balanced pattern: in some runs, multi-label slightly outperformed binary (e.g., seed 42 at 0.001: 0.794 vs. 0.692), while in others binary had a small edge (e.g., seed 123 at 0.010: 0.746 vs. 0.748). Overall, both versions fell in a similar range, with neither consistently superior.

### Bias Assessment and Cut-point determination

The best performing binary model (HuBERT, 4c) showed good performance in male (AUC 0.91, MSD present in 35/55), female (AUC 0.97, MSD present in 27/57), younger than 65 (AUC 0.97, MSD present in 30/55) and older than 65 (AUC 0.92, MSD present in 32/57) slices of the data. Performance was slightly lower for males above 65 (AUC 0.85, MSD present in 13/24) but remained high for other intersections: males below 65 (AUC 0.94, MSD present in 22/31); females above 65 (AUC 0.96, MSD present in 19/33); and females below 65 (AUC 1.0, MSD present in 8/24). For the binary model the ROC cut-point had higher sensitivity than the *z*-score one (0.871 vs 0.775, *p* = 0.005), lower specificity (0.86 vs 0.94, *p* = 0.037), and similar balanced accuracy (0.865 vs 0.857, *p* = 0.738). The ROC cut point was chosen based on superior sensitivity.

Detailed results of the multilabel cut point comparisons can be found in Table 1 in the Supplemental Data. The ROC-based cut point was numerically superior on more measures for flaccid, ataxic, and hypokinetic dysarthrias, whereas the z-score threshold was best for hyperkinetic dysarthria and apraxia. Results for spastic dysarthria were mixed, with greater sensitivity for the *z*-score threshold and greater specificity for the ROC threshold. Based on visual inspection of the cut-points in relation to the predicted probability densities (see Figure 4), it appeared that the *z*-score threshold for spastic dysarthria effectively acts as a ‘control vs MSD’ classifier, whereas the ROC threshold sacrifices sensitivity to gain performance in the ‘spastic vs other MSD’ contrast. Given our focus on sensitivity, we went with the z-score threshold, but this example demonstrates the challenge of defining a cut point that separates the MSD of interest from controls *and* from other MSDs. For instance, the other MSD densities overlap considerably with that of flaccid, spastic, and apraxia. In the case of ataxic and hypokinetic there is significant overlap between all three distributions. In contrast, it is easy to separate controls from flaccid, spastic, and apraxia.

**Figure 4.**
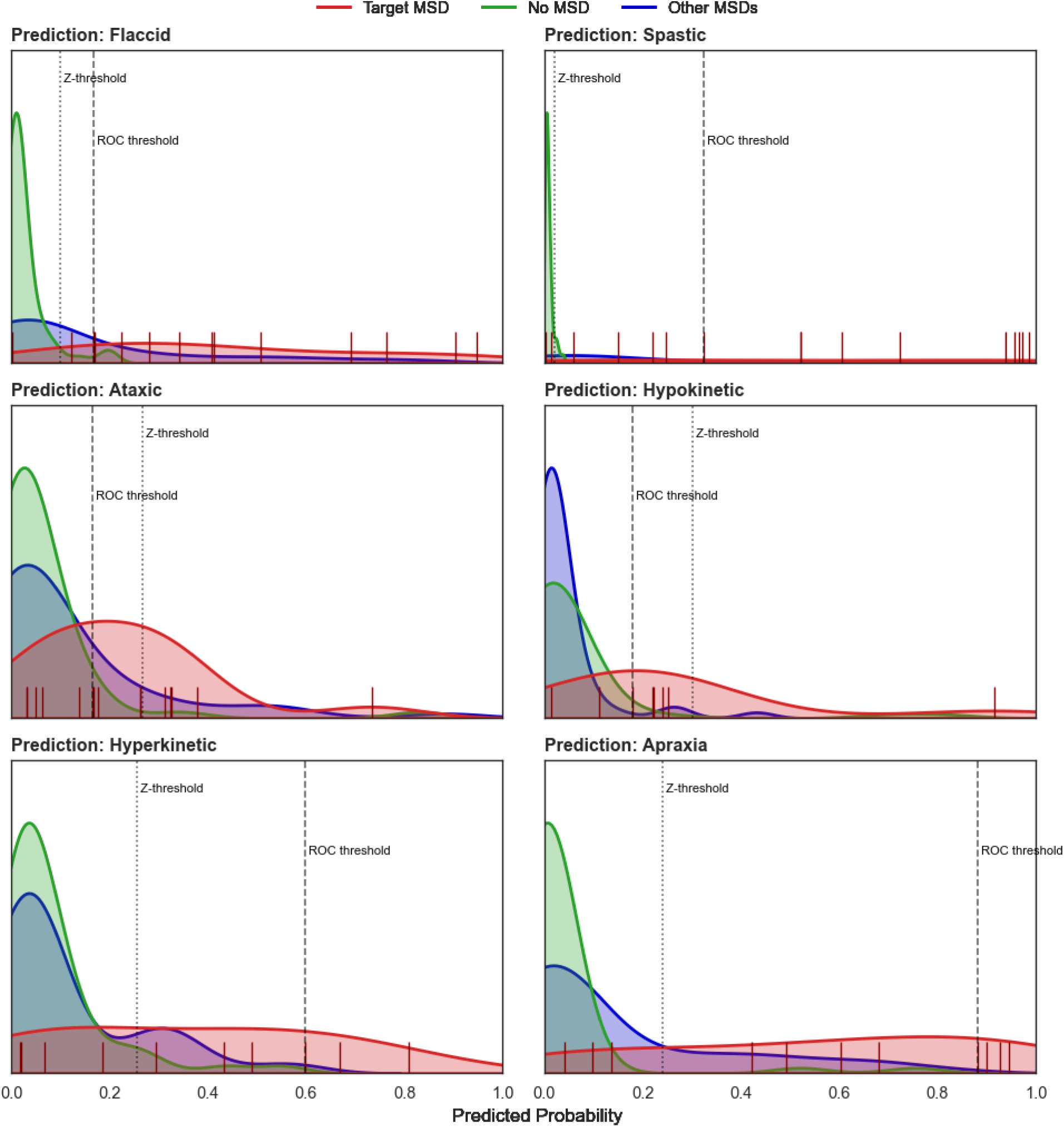
Predicted probabilities for multilabel classification on the validation set using the final multiclass model, with dotted and dashed lines indicating z- and ROC-based thresholds, respectively.

### Model Testing in Independent Datasets

We then applied the cut-points to the test set (see Table 2), where performance was maintained in the case of the binary model, which showed very high sensitivity, specificity, and balanced accuracy. For the multilabel model, AUCs in the test set ranged from moderate (0.668 in the case of ataxic dysarthria) to excellent (0.886 in the case of apraxia of speech), which suggests that the model did learn generalizable MSD features. However, AUC is a rank-based metric and we were interested in the generalization of the cut points established in the validation set. In contrast to the binary case, performance for the multilabel model deteriorated considerably. Balanced accuracy for flaccid, ataxic, hypokinetic, and hyperkinetic dysarthrias was no better than chance, with very poor sensitivity (flaccid, hypokinetic, hyperkinetic dysarthrias) or specificity (ataxic dysarthria). As expected based on Figure 4, the spastic dysarthria cut point identified all such cases at the cost of specificity, but was above chance on balanced accuracy. Apraxia of speech was the only MSD where the cut points performed reasonably well with a high balanced accuracy and specificity, but only moderate sensitivity.

**Table 2.** Performance metrics (sensitivity, specificity, balanced accuracy, and AUC) for the best binary and multilabel models evaluated on the validation and test sets using the determined cut-points.

|  | Binary (4c) | Multilabel (3c_2) |  |  |  |  |  |
| --- | --- | --- | --- | --- | --- | --- | --- |
|  | MSD | Flaccid | Spastic | Ataxic | Hypokinetic | Hyperkinetic | Apraxia |
| **Validation** |  |  |  |  |  |  |  |
| Sensitivity | 0.871 (0.790-0.952) | 0.857 (0.741-0.955) | 0.875 (0.688-1.0) | 0.667 (0.4-0.867) | 0.625 (0.25-0.875) | 0.6 (0.3-0.9) | 0.727 (0.454-1.0) |
| Specificity | 0.86 (0.76-0.94) | 0.847 (0.776-0.918) | 0.646 (0.552-0.740) | 0.866 (0.794-0.928) | 0.942 (0.894-0.981) | 0.853 (0.784-0.922) | 0.861 (0.792-0.931) |
| Balanced Accuracy | 0.865 (0.801-0.926) | 0.852 (0.745-0.939) | 0.760 – (0.656-0.844) | 0.766 (0.635-0.882) | 0.784 (0.606-0.933) | 0.726 (0.572-0.881) | 0.794 (0.648-0.926) |
| **Test (vs all)** |  |  |  |  |  |  |  |
| AUC | 0.965 (0.933-0.990) | 0.704 (0.567-0.827) | 0.875 (0.814-0.927) | 0.668 (0.533-0.792) | 0.805 (0.699-0.893) | 0.713 (0.578-0.836) | 0.886 (0.817-0.944) |
| Sensitivity | 0.936 (0.883-0.979) | 0.429 (0.143-0.714) | 1.0 (1.0-1.0) | 0.704 (0.519-0.852) | 0.158 (0.0-0.316) | 0.083 (0.0-0.25) | 0.688 (0.531-0.844) |
| Specificity | 0.967 (0.917-1.0) | 0.769 (0.694-0.837) | 0.348 (0.267-0.430) | 0.417 (0.331-0.504) | 0.985 (0.963-1.0) | 0.993 (0.979-1.0) | 0.918 (0.869-0.959) |
| Balanced Accuracy | 0.951 (0.917-0.981) | 0.599 (0.422-0.782) | 0.674 (0.633-0.715) | 0.561 (0.463-0.653) | 0.572 (0.496-0.654) | 0.538 (0.489-0.625) | 0.803 (0.717-0.882) |

To understand the misclassifications better we created co-occurrence matrices for the multilabel model alone (Figure 5a) and a scenario where cases are first classified by the binary model, and then those with an MSD are subclassified by the multilabel model (Figure 5b). We restricted this analysis to the 132 test cases with at most one MSD, but each case can still be *predicted* to have more than one MSD (i.e., the rows won’t add up to one). For the one-stage, or multilabel only, classification approach, most MSD cases are flagged as spastic and/or flaccid, along with many controls. In contrast, very few cases are predicted to have hypokinetic or hyperkinetic dysarthria. Unsurprisingly, the two-stage classification process results in better specificity and most controls are correctly identified in this paradigm. However, the confusion between spastic and ataxic dysarthria and other MSDs remains.

**Figure 5.**
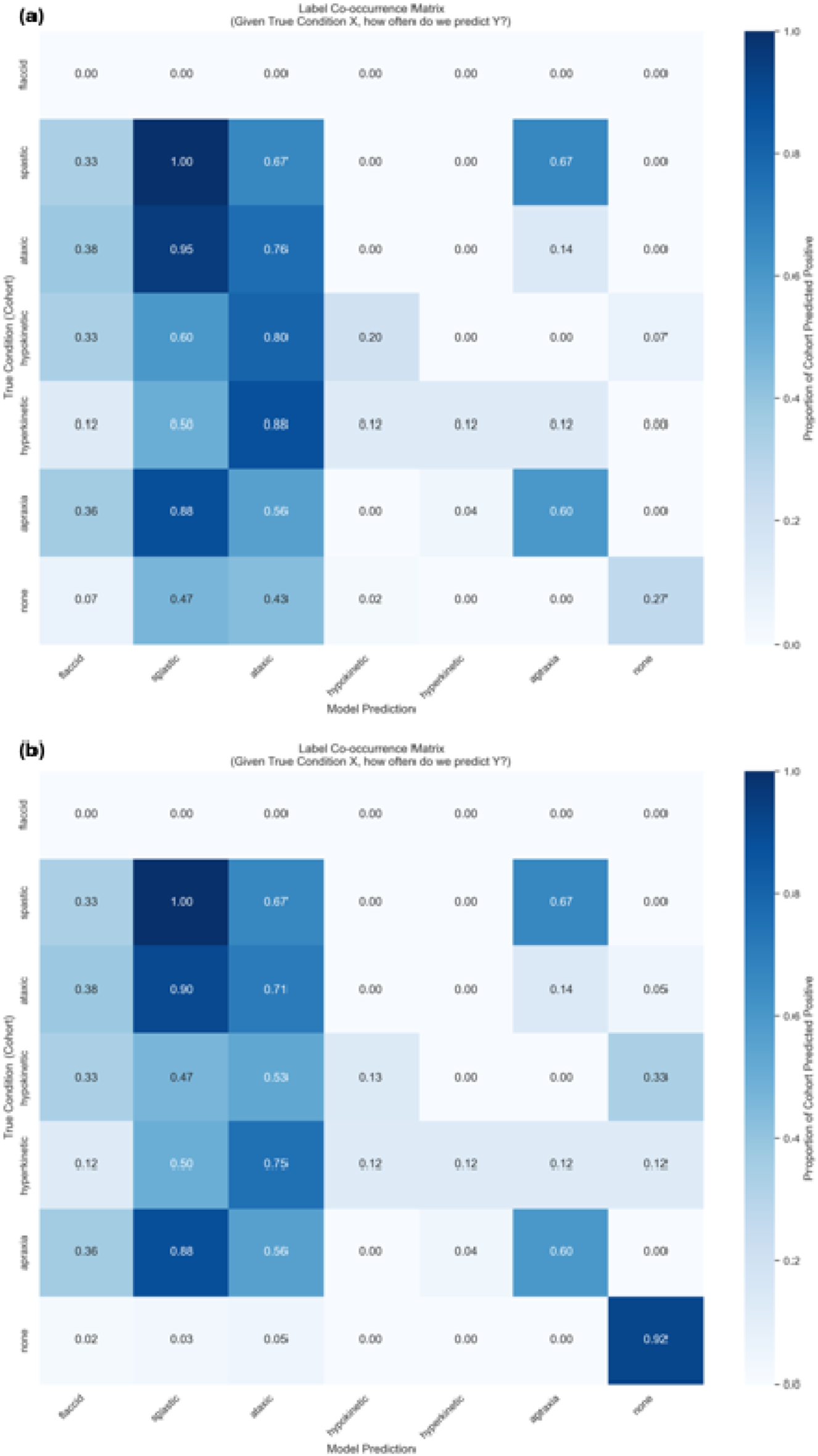
Co-occurrence matrices from standalone multilabel classification (a) and a two-step approach with initial MSD detection followed by subtype classification (b).

## Discussion

This study compared baseline models with traditional acoustic features, GRU with articulatory-informed Phonet features, and large pretrained self-supervised networks for the automatic classification of MSDs. Across both binary and multi-label tasks, Phonet- derived GRUs achieved performance that was competitive with, and in some cases surpassed, HuBERT (the strongest pretrained baseline) while consistently outperforming eGeMAPS and MFCC-based models. Subtype analyses showed that apraxia of speech and spastic dysarthria were most reliably detected, while hyperkinetic dysarthria remained the most difficult across all models.

Both self-supervised pretrained models and Phonet-derived models substantially outperformed models trained with traditional static acoustic features such as eGeMAPS and MFCCs. While these static features capture spectral and prosodic information, they do not reflect the complex temporal and articulatory patterns that characterize MSDs (Eyben et al., 2016). Phonet, in contrast, is trained to predict phonemes and phonological classes, producing features that mirror the structure of human articulation (Vásquez-Correa et al., 2019). This design is particularly valuable in MSDs, where impairments originate from disruptions in motor planning and execution rather than from abstract acoustic patterns (Ballard et al., 2015; Duffy, 2020). The strong and stable performance of phoneme embeddings (models 3c_1 and 3c_2) across subtypes further underscores the value of articulatory-informed features for clinical classification. Large pretrained models such as HuBERT and SSAST are also powerful but optimized for general-purpose tasks like speech recognition and speaker identification (Gong et al., 2022; Hsu et al., 2021), with embeddings that capture broad acoustic information.

Our results showed that pretrained and Phonet-derived models offer complementary advantages: HuBERT achieved the single highest AUC in the binary task, while Phonet- derived GRUs delivered nearly equivalent accuracy while being more compact and interpretable, underscoring the clinical potential of both model types. Distinct strengths were also evident in subtype-level analyses. Phonet-derived GRUs, especially model 3c_2, performed best for apraxia of speech, hypokinetic dysarthria, and spastic dysarthria, subtypes characterized by clear articulatory and phonatory markers. These consistent patterns align closely with Phonet’s articulatory-informed representations.

HuBERT also achieved near-ceiling accuracy for apraxia and spastic dysarthria but was weaker for hypokinetic dysarthria, likely because its representations emphasize global acoustic context rather than detailed articulatory control or the hallmark respiratory- phonatory features of hypokinetic dysarthria. SSAST showed a similar trend, excelling in apraxia of speech and spastic dysarthria but struggling with hypokinetic dysarthria.

Pretrained models such as HuBERT and SSAST may be advantageous for subtypes with strong global acoustic cues (e.g., slow speech rate, reduced pitch/loudness variation) like spastic dysarthria, while Phonet-derived features are better suited for subtypes with disproportionately prominent articulatory impairments such as apraxia of speech. This suggests that ensemble approaches combining Phonet’s articulatory grounding with pretrained models that are sensitive to more global features may yield even stronger performance across subtypes.

Some subtypes proved challenging to detect even in the training and validation sets. Hyperkinetic dysarthria proved the most difficult, perhaps because its inconsistent articulation errors, and variable voice features and prosody create highly inconsistent acoustic signals (Enderby, 2013). Ataxic dysarthria was also challenging, possibly as its hallmark irregular articulatory breakdowns are also variable (Kent et al., 2000). This study utilized a single repeated sentence, but longer and/or more natural speech samples might be more sensitive to less predictable fluctuations. The findings highlight complementary strengths of self-supervised and articulatory-informed approaches.

A key innovation of this study lies in its evaluation of hard classification decisions on a fully independent test set, moving beyond standard reporting of ranking metrics. While most prior work in this domain focuses on internal validation or model discrimination, few studies assess whether models can make reliable, threshold-based decisions that translate to unseen clinical data. This step is critical for real-world implementation, where models must not only identify patterns but also support actionable outcomes such as diagnosis, triage, or referral. The best-performing binary model (i.e., model 4c) achieved high accuracy in distinguishing individuals with and without MSDs, with strong validation performance that generalized well to the test set. Sensitivity and specificity remained above 0.93, and the threshold defined in the validation set maintained balanced accuracy in independent test datasets, supporting its potential for clinical screening. In contrast, the best multi-label model (i.e., model 3c_2) demonstrated clear subtype-specific learning, with strong area under the curve values across subtypes and consistent detection of apraxia and spastic dysarthria in the test set. However, the decision thresholds did not translate well. Sensitivity and specificity dropped for several subtypes, highlighting the challenge of threshold calibration in small and heterogeneous datasets and the difficulty of separating overlapping speech patterns across MSD subtypes.

It is worth noting that because this was a more realistic test scenario than typically used in the literature, it was likely made more challenging by the fact that the sentence repetitions used were not in response to the standardized prompt used in the training and validation sets, since they were obtained under entirely different circumstances spanning several decades. The small validation set also limited our ability to calibrate thresholds, likely contributing to reduced sensitivity and specificity for some subtypes in the test set. This challenge is common in rare neurological diseases and underscores the need for calibration methods that perform reliably under data-limited conditions.

Nevertheless, our findings suggest that while advanced models can accurately detect and differentiate MSDs, further work is needed to ensure reliable thresholding for subtype classification in clinical practice.

In conclusion, this study shows that advanced modeling approaches, particularly self-supervised pretrained models and articulatory-informed representations, substantially outperform traditional static acoustic features for detection of MSDs. Binary classification of MSD presence achieved excellent performance and generalized well to independent datasets, including threshold-based decisions, which is essential for real-world implementation. In contrast, multi-label classification revealed promising subtype-specific learning but poor generalization of cut-points, underscoring the challenge of calibration in rare neurological disorders. Subtype confusion was common, reflecting the clinical reality of overlapping MSDs and highlighting the complexity of categorical classification. By evaluating these hard calls and the threshold stability on independent data, this work addresses a critical gap in the literature and moves toward clinically actionable AI-informed tools. Future research should prioritize robust calibration strategies, larger and more diverse datasets, and integration into clinical workflows to evaluate practical deployment.

## Acknowledgement

The authors thank Ashley D. Bachman for project administration and also thank the patients and caregivers who have generously committed themselves to this research.

---

**Figure 6.** Binary classification performance across models. Bars show median validation AUCs, and error bars indicate the range (minimum to maximum) across seeds.

**Figure 7.** Multi-label classification performance across models. Bars show median validation AUCs, and error bars indicate the range (minimum to maximum) across seeds.

**Figure 8.** Subtype-specific validation AUCs for the best-performing configuration of each model family in the multi-label classification task.

**Figure 9.** Predicted probabilities for multilabel classification on the validation set using the final multiclass model, with dotted and dashed lines indicating z- and ROC-based thresholds, respectively.

**Figure 10.** Co-occurrence matrices from standalone multilabel classification (a) and a two-step approach with initial MSD detection followed by subtype classification (b).

**Table 3.** Distribution of MSD Subtypes in the Training, Validation, and Test Datasets.

**Table 4.** Performance metrics (sensitivity, specificity, balanced accuracy, and AUC) for the best binary and multilabel models evaluated on the validation and test sets using the determined cut-points.

