## Supplemental for "Automated Detection of Motor Speech Disorders and Subtype Classification"

### Supplemental Data

#### The Two Independent Test Datasets

The first (77 participants/recordings) was taken from the Neurodegenerative Research Group (NRG), which recruits neurodegenerative speech and language disorders into several NIH funded projects. Patients enrolled in NRG are assigned an MSD subtype based on a comprehensive in-person speech language battery with consensus across two board certified SLPs. The second (77 participants/recordings) was taken from the archives of one of the authors (J.R.D), acquired during evaluation of dysarthria in the 1990s and 2000s.

#### Model Training Procedures

Training procedures were tailored to each model family. Across all model families, the same data splits and seed protocols were maintained to ensure comparability. Logistic regression baselines (models 1a and 1b) were trained with an L2 regularization penalty, and model strength was optimized by searching across a wide range of regularization values (from very strong to very weak). GRU-based models (models 2a, 2b, and 3a–3c) used bidirectional gated recurrent units with 128 hidden dimensions, one recurrent layer, dropout (0.3), layer normalization, and attention pooling to capture temporal structure. These models were trained with the Adam optimizer (weight decay = 0.01) using binary cross-entropy loss for both binary and multi-label tasks.

The pretrained self-supervised models HuBERT (models 4a–4c) and SSAST (models 4d–4f) were evaluated under three conditions: using frozen encoders with only a classifier trained (models 4a and 4d), partially fine-tuning the top encoder layers (models 4b and 4e), or fully fine-tuning all model parameters (models 4c and 4f). Optimization used the AdamW algorithm with weight decay (0.01), gradient accumulation across four mini-batches, and adaptive learning rate reduction (by half) if validation performance failed to improve for two consecutive epochs.

To assess robustness, all GRU-based, HuBERT, and SSAST models were trained under three random seeds (42, 123, and 2025) and three learning rates (0.0001, 0.001, 0.01) per seed, for up to 100 epochs with early stopping after five consecutive epochs without improvement.

All models were implemented in Python using a combination of standard machine learning and deep learning libraries. For the baseline models, we relied on scikit-learn with preprocessing steps such as standardization. Acoustic features were extracted with openSMILE for eGeMAPS and with librosa for MFCCs. For the GRU-based, HuBERT, and

SSAST models, we used PyTorch for model construction, optimization, and training. Phonet-based features were extracted using the Phonet toolkit.

#### Model Evaluation and Selection

To ensure robust comparisons across model families, final model selection was based on the learning rate that maximized the minimum validation AUC (binary) or the minimum macro-averaged AUC (multi-label) across seeds. This conservative strategy ensured that results reflected configurations performing reliably across different initializations, rather than those that excelled only under favorable conditions. In addition, for multi-label classification, we reported the single best-performing model (highest macro-averaged validation AUC) to examine subtype-level performance. To further evaluate whether the multi-label approach improved subtype classification, we also trained binary versions of models 3c_2, 4c, and 4f specifically on hyperkinetic dysarthria, as this was the most challenging subtype in the multi-label setting, and compared their performance with the corresponding multi-label models across seeds and learning rates.

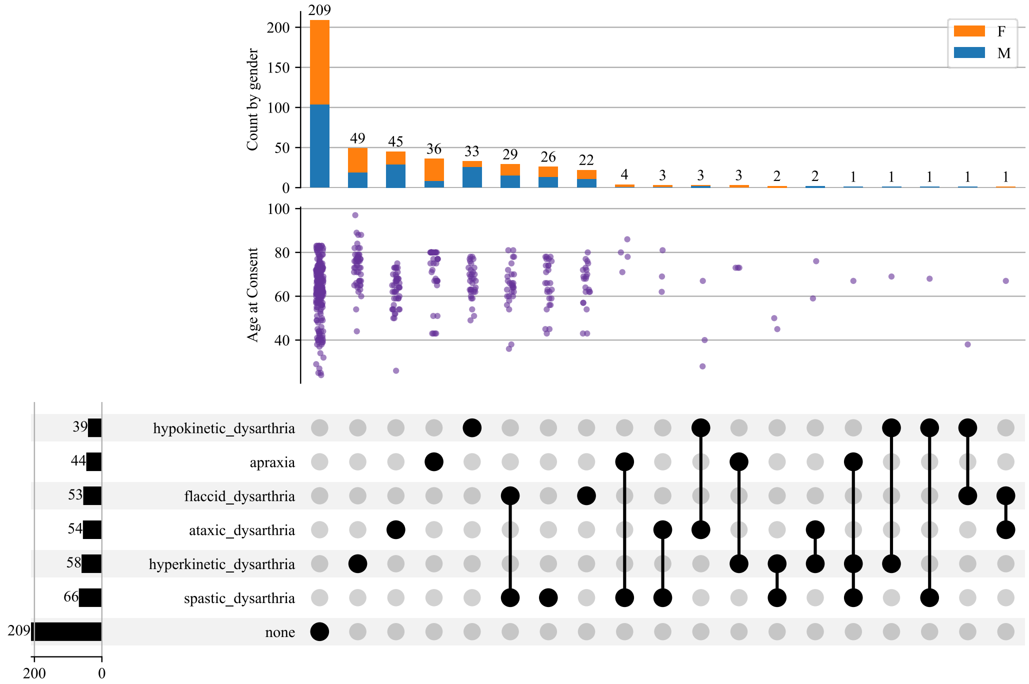


*Figure 1. Breakdown of training recordings by combination of MSDs along with age and gender.*

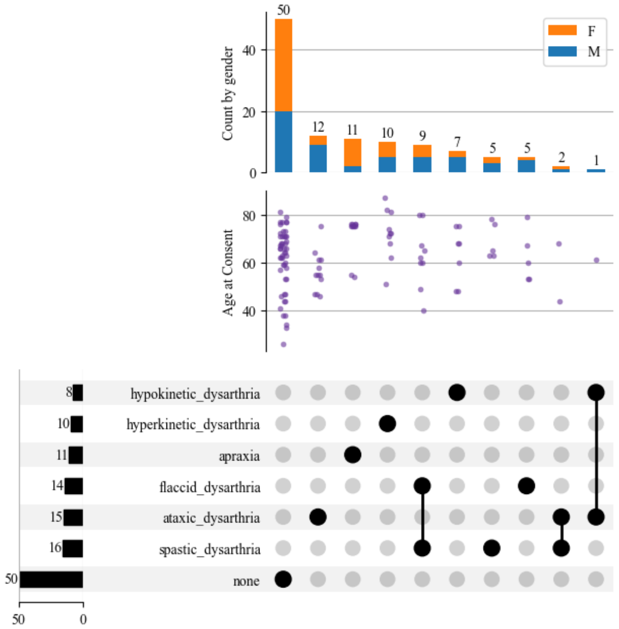


*Figure 2. Breakdown of validation recordings by combination of MSDs along with age and gender.*

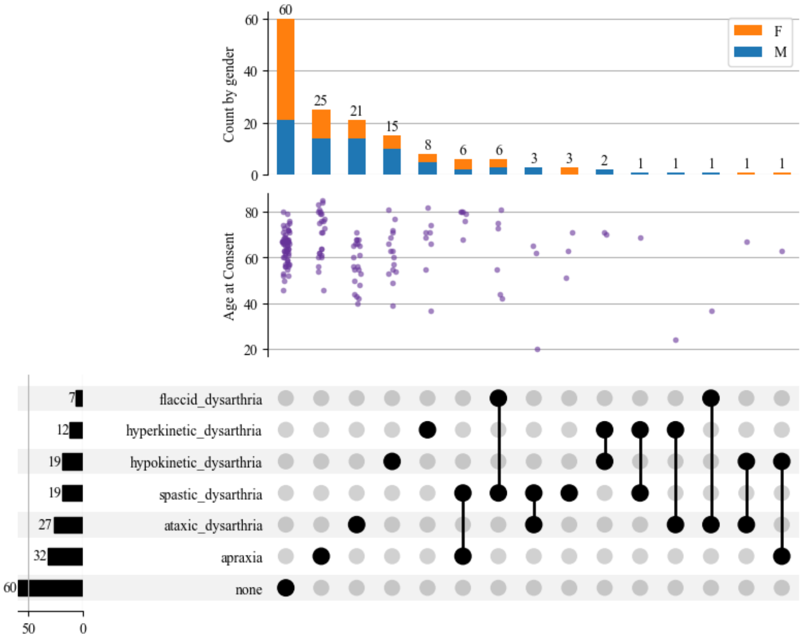


*Figure 3. Breakdown of test recordings by combination of MSDs along with age and gender.*

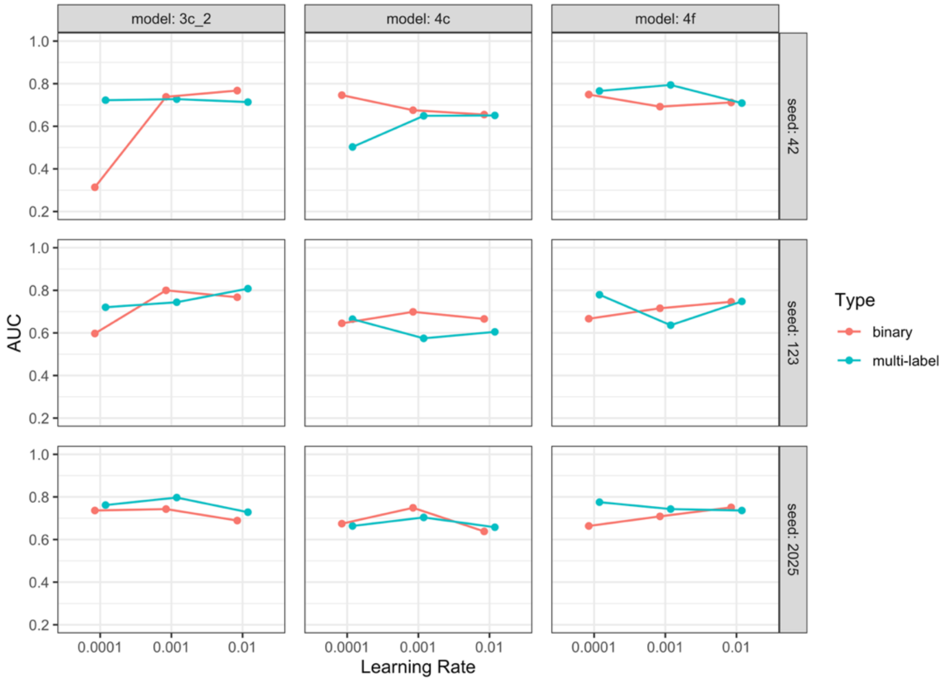


*Figure 4. Binary vs. multi-label classification performance for hyperkinetic dysarthria using models 3c_2, 4c, and 4f across seeds and learning rates.*

*Table 1. Comparison of diagnostic performance between ROC-derived and Z-score-based cut-points across motor speech disorder subtypes. Point estimates are reported for each method. The difference in performance (Delta) and its 95% confidence interval were calculated using a stratified paired bootstrap procedure (n=2,000).*

| **Diagnosis** | **Metric** | **ROC (Est)** | **Z-Score (Est)** | **Difference (95% CI)** | ***P*-Value** |
|---|---|---|---|---|---|
| **Flaccid Dysarthria** | Sensitivity | 0.857 | 0.929 | -0.071 (-0.214, 0.000) | 0.698 |
| | Specificity | 0.847 | 0.796 | 0.051 (0.010, 0.102) | 0.01 |
| | Balanced Accuracy | 0.852 | 0.862 | -0.010 (-0.092, 0.041) | 0.865 |
| **Spastic Dysarthria** | Sensitivity | 0.563 | 0.875 | -0.313 (-0.562, -0.125) | 0.002 |
| | Specificity | 0.927 | 0.646 | 0.281 (0.198, 0.375) | <0.001 |
| | Balanced Accuracy | 0.745 | 0.76 | -0.016 (-0.141, 0.099) | 0.867 |
| **Ataxic Dysarthria** | Sensitivity | 0.667 | 0.333 | 0.333 (0.133, 0.600) | 0.006 |
| | Specificity | 0.866 | 0.918 | -0.052 (-0.103, -0.010) | 0.01 |
| | Balanced Accuracy | 0.766 | 0.625 | 0.141 (0.025, 0.274) | 0.012 |
| **Hypokinetic Dysarthria** | Sensitivity | 0.625 | 0.125 | 0.500 (0.125, 0.875) | 0.005 |
| | Specificity | 0.942 | 0.971 | -0.029 (-0.067, 0.000) | 0.088 |
| | Balanced Accuracy | 0.784 | 0.548 | 0.236 (0.053, 0.423) | 0.005 |
| **Hyperkinetic Dysarthria** | Sensitivity | 0.2 | 0.6 | -0.400 (-0.700, -0.100) | 0.013 |

| | | | | | |
|---|---|---|---|---|---|
| | Specificity | 1 | 0.853 | 0.147 (0.078, 0.216) | <0.001 |
| | Balanced Accuracy | 0.6 | 0.726 | -0.126 (-0.291, 0.024) | 0.092 |
| **Apraxia** | Sensitivity | 0.273 | 0.727 | -0.455 (-0.727, -0.182) | 0.002 |
| | Specificity | 1 | 0.861 | 0.139 (0.069, 0.208) | <0.001 |
| | Balanced Accuracy | 0.636 | 0.794 | -0.158 (-0.314, -0.012) | 0.033 |
